# Blood-derived DNA methylation clusters associate with adverse social exposures and endophenotypes of stress-related psychiatric illness in a trauma-exposed cohort of women

**DOI:** 10.1101/2022.03.24.22272373

**Authors:** JR Pfeiffer, Sanne J.H. van Rooij, Yara Mekawi, Negar Fani, Tanja Jovanovic, Vasiliki Michopoulos, Alicia K. Smith, Jennifer S. Stevens, Monica Uddin

## Abstract

Adverse social exposures (ASEs) such as low income, low educational attainment, and childhood/adult trauma exposure are associated with variability in brain region measurements of grey matter volume (GMV), surface area (SA), and cortical thickness (CT). These CNS morphometries are associated with stress-related psychiatric illnesses and represent endophenotypes of stress-related psychiatric illness development. Epigenetic mechanisms, such as 5-methyl-cytosine (5mC), may contribute to the biological embedding of the environment but are understudied and not well understood. How 5mC relates to CNS endophenotypes of psychiatric illness is also unclear.

In 97 female, African American, trauma-exposed participants from the Grady Trauma Project, we examined the associations of childhood trauma burden (CTQ), adult trauma burden, low income and low education with blood-derived 5mC clusters and variability in brain region measurements in the amygdala, hippocampus and frontal cortex subregions. To elucidate whether peripheral 5mC indexes CNS endophenotypes of psychiatric illness, we tested whether 73 brain/blood correlated 5mC clusters, defined by networks of correlated 5mC probes measured on Illumina’s HumanMethylation Epic Beadchip, mediated the relationship between ASEs and brain measurements.

CTQ was negatively associated with rostral middle frontal gyrus (RMFG) SA (β = - 0.231, p = 0.041). Low income and low education were also associated with SA or CT in a number of brain regions. Seven 5mC clusters were associated with CTQ (pmin = 0.002), two with low education (pmin = 0.010), and three with low income (pmin = 0.007). Two clusters fully mediated the relation between CTQ and RMFG SA, accounting for 47% and 35% of variability respectively. These clusters were enriched for probes falling in DNA regulatory regions, as well as signal transduction and immune signaling gene ontology functions. Methylome-network analyses showed enrichment of macrophage migration (p = 9×10^-8^), T cell receptor complex (p = 6×10^-6^), and chemokine-mediated signaling (p = 7×10^-4^) pathway enrichment in association with CTQ.

Our results support prior work highlighting brain region variability associated with ASEs, while informing a peripheral inflammation-based epigenetic mechanism of biological embedding of such exposures. These findings could also serve to potentiate increased investigation of understudied populations at elevated risk for stress-related psychiatric illness development.

## Introduction

Adverse social exposures (ASEs) such as low income, low educational attainment, childhood trauma, and adult trauma exposure, are common in the United States (US). Over the past five years, poverty rates in the US have ranged from 12 to 15%[1], and are likely to increase due to the impact of the COVID-19 pandemic[2]. The percentage of US adults aged 18 to 24 that finish high school with a diploma or GED is roughly 88%, meaning that over one million students per year leave high school without completion credentials[3]. Past year exposure to physical abuse in US children ranges from 4% to 16%, whereas past year psychological abuse is reported in 10% of children in the US; sexual abuse is experienced at some point during childhood in 15% to 30% of children[4]. Furthermore, in a US population-based study of 34,653 participants, researchers found that depending on self-reported race/ethnicity, respondents’ prevalence of lifetime trauma exposure varied between 66% and 84%[5]. Other researchers have observed a lifetime traumatic exposure prevalence ranging from 50% to 90%[6-8]. ASEs are known to have long-term impact on emotional well-being and mental health; however, the mechanistic basis for these effects remain poorly understood.

One particularly salient risk following ASE exposure is the development of stress-related psychiatric illness or symptoms of such illnesses. For example, in both adults and children, poverty-related stress is associated with increased internalizing symptoms (e.g., sadness or anxiety), involuntary engagement stress responses (e.g., emotional arousal or intrusive thoughts), externalizing symptoms (e.g., acts of aggression or theft), and involuntary disengagement stress responses (e.g., emotional numbing or cognitive interference) [9]. Relatedly, World Health Organization (WHO) data has shown that rates of anxiety, mood, and substance-use disorders are all positively correlated with measures of socioeconomic disadvantage, including both low income and low educational attainment [10]. Moving a step beyond correlation, a longitudinal study found that higher educational attainment significantly attenuated both the overall risk of adult onset major depressive disorder (MDD), particularly in women, and the conditional risk of MDD as well[11].

The effects of childhood maltreatment or trauma on long-term mental health prognoses are also well-documented. In a recent meta-analysis of over 190 studies and 68,000 participants, researchers observed that higher childhood maltreatment (including potential traumas such as emotional abuse, physical abuse, sexual abuse) scores were associated with MDD diagnosis later in life and with depression symptom severity[12]. This built upon previous work showing that exposure to any form of maltreatment during childhood was associated with a two-fold risk of MDD development, and nearly a three-fold risk of generalized anxiety disorder (GAD) development in adulthood[13]. Although childhood and adolescence are regarded as particularly vulnerable periods of development, the impact of trauma on psychiatric illness/symptom development are also notable when trauma is experienced in adulthood, with effects that are known to vary by race/ethnicity and gender. A particularly salient example of this is evident in the potential development of post-traumatic stress disorder (PTSD) after trauma. Recent population-based work highlights race/ethnic differences in psychiatric illness development and PTSD in particular. Researchers found that African Americans are generally more resilient to developing psychiatric illness after a TE, compared to other races/ethnicities[14]; however, this resilience does not apply to PTSD, where African Americans experience elevated conditional PTSD development rates relative to other races/ethnicities[14]. This, in tandem with research showing that females are subject to almost double the risk of developing PTSD and other mood-anxiety disorders throughout their lifetime[6, 15-17], highlights that African American women in particular may be subject to elevated risk of stress-related psychiatric illness development.

A great deal of work has been placed in identifying the neural correlates of poverty, low education, and childhood/adult trauma exposure. Researchers have identified specific signatures of central nervous system (CNS) variability in association with ASEs using methods such as structural magnetic resonance imaging (MRI) and functional MRI (fMRI). In rodent [18, 19] and human [20-24] studies, observed neural correlates of ASEs include variability in volumetric, cortical thickness (CT), surface area (SA), excitability, and connectivity measures. Brain regions including frontal cortex, hippocampus, and amygdala, contribute to adaptive behavioral and emotional responses, and are relevant to psychiatric illness development due to their contributions to the generation and regulation of major decision-making, memory formation, and stress-reactivity processes [25]. To this end, a meta-analytic study showed that frontal pole (FP), superior frontal gyrus (SFG), and rostral middle frontal gyrus (RMFG) frontal cortex subregions are heavily involved in attention, working memory, cognitive flexibility, and executive reasoning [26], with added involvement of the frontal pole in introspection and longitudinal thinking [27]. The anterior cingulate cortex (ACC) is regarded as a mediator of the interactions between higher level cognitive processes and emotion/stress reactivity[28], and the orbito-frontal cortex (OFC) contributes to decision making and emotional processing functions[29]. These functions are impaired in numerous mental illnesses, including but not limited to PTSD[30], MDD[31], GAD[32], bipolar disorder[33], and schizophrenia[33]. These neural correlates of ASEs are associated with the development of stress-related psychiatric illnesses and may represent neural endophenotypes of said disorders[34-36].

Importantly though, the effects of ASEs do not travel directly from environment to psychiatric illness. The cascade of effect, rather, travel initially through the neuroendocrine system as a response to stressful external stimuli[37], whereby cortisol signaling occurs throughout the body, engaging immune (and other) biological networks[38]. This immune network activation and responses to cortisol signaling result in neural growth, inflammation, metabolism, and stress-related pathway disruption through epigenetic and other molecular mechanisms[39, 40]. The process of DNA methylation, which commonly refers to the addition of a 5-methyl-cytosine (5mC) residue to a cytosine-phosphate-guanine (CpG) base-pair, is one such epigenetic mechanism, and is understood to enhance or reduce mRNA transcription of genes in an experience-dependent, and temporally-stable manner[41]; continued research has pointed to the importance of 5mC in this biological embedding of external stimuli[42, 43]. Because of challenges in taking epigenetic measurements from a living human brain, the primary etiologic tissue of interest in regard to psychiatric illness development, peripheral tissues such as blood or saliva serve as proxies for etiological tissue. It is well-documented that these peripheral epigenetic measures can index changes to the hypothalamic-pituitary-adrenal (HPA)-axis [44, 45], immune system [46, 47], and CNS [48-50], although the direction of effect may be discordant across tissues [49, 51]. Stress-related physiological responses and their accompanying epigenetic alterations are thought to confer the biological embedding of ASEs [52, 53], and can ultimately increase the risk of stress-related psychiatric illness [39, 54]. Peripheral epigenetic measures can also index CNS-relevant endophenotypes of psychiatric illness development, as observed in studies using neuroimaging and peripheral epigenetic measures in tandem. However, these studied have primarily utilized candidate gene approaches, investigating 5mC in the *SLC6A4* [55-57], *NR3C1* [58, 59]], *FKBP5* [60], and *SKA2* [61, 62] genes, and neuroimaging measures of structure and function from the frontal cortex, hippocampus, and amygdala. Findings show that peripheral 5mC can index CNS structural variability[55-62] associated with psychiatric illness development, and that locus-specific peripheral 5mC can mediate ASE-associated CNS structure variability[62].

To date, however, limited work has investigated the relationships among ASEs, peripheral 5mC, and CNS endophenotypes of psychiatric illness at a genome-scale. To this end, the current exploratory study applied genome-scale approaches in an all-female, African American sample from the Grady Trauma Project (GTP) to assess whether blood-derived 5mC measurements might index CNS endophenotypes of stress-related psychiatric illness. We focus specifically on the GTP because African American women may be subject to elevated risk of stress-related psychiatric illness development, but the mechanisms underlying that elevated risk are still unclear. In this study, we were specifically interested whether clusters of peripheral 5mC measurements statistically mediate the relation between ASEs and fronto-limbic grey matter volume (GMV), SA, and CT measures. Based on previous work, we hypothesized that identified 5mC modules would be enriched with 5mC probes falling in genes with HPA-axis[44, 45], immune system[46, 47], and CNS-relevant [48-50] gene ontology (GO) functions.

## Materials and Methods

### Participants

The current research draws on 97 female, trauma-exposed participants. Participants were part of the GTP, a larger investigation of genetic and environmental factors that predict the response to stressful life events (SLEs) in a predominantly African American, low-income, urban population [15]. Research participants were approached in the waiting rooms of primary care clinics of a large, public hospital. After the subjects provided written informed consent, they participated in a verbal interview and gave a blood sample for genetic and epigenetic analyses. Participants in this cohort also had available demographic, psychosocial, and neuroimaging data. Exclusion criteria included intellectual disability or active psychosis. Participants provided written, informed consent for all parts of the study, and the Institutional Review Boards of Emory University and Grady Memorial Hospital approved the study procedures.

### Exposures of interest

Income, education, adult trauma burden, and childhood trauma burden were considered exposures of interest in current analyses. *Income:* Self-reported approximate household monthly income (“household income”) was measured as part of a demographic-focused inventory. Participants indicated total household income within one of the following ranges: $0 to $499, $500 to $999, or greater than $1000 per month. *Education:* Participants also reported educational attainment history (“education”) and were binned into one of the following categories: did not complete high school, completed high school or GED, or completed more schooling than high school. *Traumatic events inventory:* Adult trauma burden was assessed via a semi-structured interview using the Traumatic Events Inventory (TEI), a scale developed by the GTP researchers during their prior work in the Grady hospital primary care population [63]. TEI responses pertaining to childhood were excluded in favor of the Childhood Trauma Questionnaire (CTQ). In the current study, the continuous TEI score was used (“TEI”); the higher the TEI score, the more traumatic life events one encountered. *Childhood trauma questionnaire:* Childhood trauma burden was assessed using the self-report 28 question CTQ [64]. This inventory consists of five subscales, including sexual abuse, physical abuse, emotional abuse, as well as physical and emotional neglect. In the current study, the total CTQ score was used (“CTQ total”). Information regarding previous usage of exposure of interest variables in GTP studies can be found elsewhere [15, 42, 65-68].

*Neuroimaging* Structural MRI study procedures followed the same methods as previously described in the GTP cohort [69-72]. Structural images were acquired on a Siemens 3.0-Tesla Magnetom Trio TIM whole-body MR scanner (Siemens, Malvern, PA, USA) with a 12-channel head coil, and using a gradient-echo, T1-weighted pulse sequence (176 slices, TR = 2600 ms, TE = 3.02 ms, 1 mm3 voxel size). The T1 scan was processed using FreeSurfer (available at http://surfer.nmr.mgh.harvard.edu), and a standardized protocol (available at http://enigma.ini.usc.edu/protocols/imaging-protocols/) was used to check the quality of the segmentations before further analyses were performed. Hemisphere-specific frontal cortex CT and SA measurements were taken along with amygdala and hippocampal gray matter volume to assess brain morphometry of the fronto-limbic pathway. For the purpose of the current study, within each brain region (hippocampus and amygdala), we took the mean volume of the two hemispheres (e.g., hemisphere-mean hippocampal volume, hemisphere-mean amygdala volume). CT and SA develop along discordant temporal trajectories[73], and are affected in different manners based on underlying genetic sequence[74]. Based on this rationale, we examined CT and SA measures of frontal cortex subregions, instead of gray matter volume. Frontal cortex subregions included in the current study map to Brodmann’s areas 8, 9, 10, 11, 24, 25, 32, and 46[25], and include FP, medial OFC (mOFC), lateral OFC (lOFC), SFG, RMFG, rostral ACC (rACC), and caudal ACC (cACC). For frontal cortex SA and CT, we used an average of values taken across hemispheres (e.g., hemisphere-mean RMFG SA, hemisphere-mean RMFG CT).

### Molecular

Whole-blood was collected in EDTA vacuum tubes prior to DNA extraction. Genomic DNA was extracted and purified using the AllPrep DNA/RNA Mini Kit (Qiagen, Valencia, CA) using manufacturer recommended methods. DNA was denatured and bisulfite converted (BSC) using the EZ DNA Methylation-GoldTM Kit (Zymo Research, Irvine, CA). After conversion, BSC DNA was applied to the Infinium MethylationEPIC BeadChip (Illumina; San Diego, California) (850k) using manufacturer recommended protocol to measure DNA 5mC at ∼850,000 loci.

### 5mC pre-processing

Beta-values measured from the 850k platform were background corrected with GenomeStudio, quality controlled (QCed), and filtered according to previously published methods[75]. All quality control and pre-processing was performed in R version 3.6.1[76]. These steps removed low quality and potentially cross-hybridizing probes, quantile-normalized probe Beta-values, and removed technical and batch effects[77-80]. 5mC beta-values were variance stabilized and logit-transformed into M-values[81]. X- and Y-chromosome-mapped probes were removed, along with *rs*-mapped probes. The remaining ∼827k probes were then subset to include only those with observed nominally significant Pearson correlation (p < 0.05) between blood and brain tissue from the ImageCpG data repository[82], which focused analysis onto loci with greater prospect for proxy or surrogate status with etiologically-relevant CNS tissue. Following QC and filtering steps, 92,208 probes remained.

### Covariates

Age, leukocyte cell estimates, intra-cranial volume (ICV), multi-dimensional scaling (MDS) of genomic ancestry, an epigenetic proxy of smoking[83], and current employment/disability status were included in models as covariates, where pertinent. *Leukocyte cell estimates:* DNAm measurements were used to estimate the proportions of granulocytes, monocytes, B cells, natural killer cells, CD4T and CD8T cells within each sample. Calculations were done according to methodology described in Houseman et al[84]. Estimated proportions of monocytes, B cells, natural killer cells, CD4T and CD8T were included as covariates in multiple regression models, where pertinent. *Genomic ancestry:* To avoid potential inaccuracies/confounding effects of self-reported race/ethnicity, genetic ancestry was modeled using MDS measures extracted from participant genome-wide association study (GWAS) data using PLINK[85]. The first four MDS genetic ancestry measures were chosen to be used as covariates based on visual inspection of scree plots, in line with previous work[86]. *Smoking:* Cigarette smoking is known to exert significant effects across the methylome[87]. Using a method discussed in detail elsewhere[88], the current study corrected for the methylomic effects of smoking by calculating effect size estimates of the top twenty-six probes from a recent smoking epigenome-wide association study (EWAS) performed in participants with African American genomic ancestry[83]. *Employment and disability:* As applied previously in the GTP [66], employment/disability status was considered as: unemployed; unemployed receiving disability support; or employed, with or without disability support.

### Probe clustering

To remove non-desired effects from probe M-values, we composed linear models in R using age, leukocyte cell esimates, genomic ancestry, and the smoking proxy as predictors of probe-wise 5mC M-values. For each probe, residual values (“residualized M-values”) were extracted for clustering. Taking the 92,208 residualized M-values, the “WGCNA” package was then used in R to build a co-methylation network[89]. First, scale-free topology model fit was analyzed. As recommended, a soft-threshold value of ten was chosen based on the lowest power at which adjusted R^2^ > 0.90. Adjacency and dissimilarity matrices were generated, and unsupervised hierarchical clustering was used to generate a clustered residual M-value network. Setting a minimum cluster size of ten generated 73 clusters of 5mC probes, where each cluster was identified by a unique color. After clustering, we extracted the first principal component of each cluster, referred to from here on as a “module eigengene” (ME). Compared to EWAS-type analyses, which assess differential methylation on the level of individual 5mC loci, network-based methods, as used in the current research, utilize dimension-reduction techniques to create networks of related 5mC probe clusters. This reduces the burden of multiple hypothesis testing across hundreds of thousands of probes, while enabling investigation into the biological significance of clustered probes, and provides the potential for increased statistical power in circumstances with a small number of biological replicates[90].

### Statistical analyses

To understand the relationships between variables used throughout the current analyses, we carried out Pearson correlations and mapped their correlation coefficients. We then conducted a set of multi-variate linear regression analyses, as shown in Figure 1. Figure 1 Arm A analyses included hemisphere-mean hippocampus and amygdala volumes as dependent variables (in separate models). Income, education, CTQ total, and TEI were included as independent variables of interest, whereas genomic ancestry, age, employment/disability and ICV were included as covariates. The same predictors and covariates were included, minus ICV, to predict frontal cortex subregions SA and CT measures. Overall CT and/or SA were not included as covariates based on prior work with this cohort[91]. Figure 1 Arm B analyses included 5mC MEs as dependent variables (in separate models). Income, education, CTQ total, and TEI were included as independent variables of interest, and employment/disability was included as a covariate. Covariates were limited in this stage of analysis due to the removal of confounding effects through residual extraction in prior pre-processing steps. Figure 1 Arm C analyses included individual fronto-limbic morphometry measures as dependent variables (in separate models). Individual 5mC MEs were included as independent variables of interest, whereas age, genomic ancestry, employment, and ICV (ICV was only included in hippocampus and amygdala models) were included as covariates. Within each phase of the analyses, including mediation analyses, continuous dependent and independent variables were standardized, resulting in standardized effect estimates. Nominal p-values were corrected for multiple hypothesis testing by controlling the false discovery rate (FDR=0.10) using the Benjamini Hochberg (BH) procedure(85). Briefly, for each nominal p-value, a BH critical value was calculated where nominal p-value’s assigned rank over the number of tests was multiplied by the accepted FDR. Nominal p-values less than this threshold were deemed BH-significant. Due to the exploratory nature of the current work, both nominal and BH-significant terms were considered for interpretation; and nominal p-values are presented within the results section of the current text.

**Figure 1.**
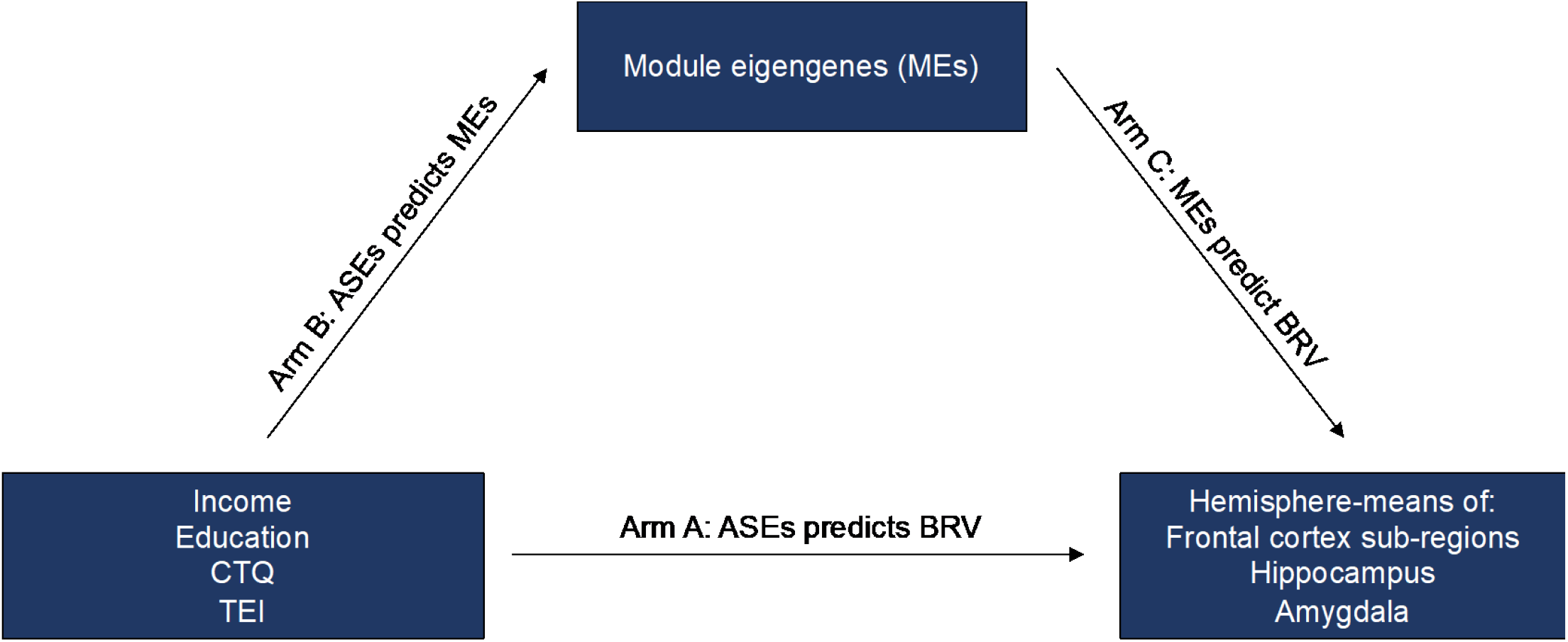
Conceptual model testing individual module eigengenes (MEs) as mediators of the hypothesized associations between adverse social exposures (ASEs) and variability in fronto-limbic brain morphometries. Arm A. Analyses included hemisphere-mean hippocampus and amygdala volumes as dependent variables (in separate models). Income, education, CTQ total, and TEI were included as independent variables of interest, whereas genomic ancestry, age, employment/disability and ICV were included as covariates. The same predictors and covariates were included, minus ICV, to predict frontal cortex subregions SA and CT measures. Arm B. Analyses included 5mC MEs as dependent variables (in separate models). Income, education, CTQ total, and TEI were included as independent variables of interest, and employment/disability was included as a covariate. Arm C. Analyses included individual fronto-limbic morphometry measures as dependent variables (in separate models). Individual 5mC MEs were included as independent variables of interest, whereas age, genomic ancestry, employment, and ICV (ICV was only included in hippocampus and amygdala models) were included as covariates.

### Mediation analyses

MEs were tested for statistical mediating status between independent variables of interest (ASEs) and dependent variables (hemisphere-mean fronto-limbic brain morphometry) using the “mediation” package in R (Figure 1). Importantly, only fronto-limbic brain morphometry measures nominally associated with income, education, CTQ total or TEI (Figure 1, Arm A) were considered. Similarly, MEs tested for mediation included only those nominally associated with an exposure (Figure 1, Arm B) and a fronto-limbic brain morphometry measure (Figure 1, Arm C). For each ME, average indirect effect (IDE), direct effect (DE), and total effect estimates and confidence intervals were calculated as a result of 10,000 quasi-Bayesian Monte Carlo approximations, applied with the mediation package. Consistent with methodology employed in the field [92, 93], we considered an ME a full mediator if the DE=0 while the IDE and total effect ≠ 0, or a partial mediator if the DE, IDE, and total effect (TE) ≠ 0. Individual probes from full mediator modules were assessed for mediation status, in order to gain insight into potential locus-specific mediation effects.

### Gene set enrichment analyses

To assess underlying methylomic network enrichment, all four ASEs of interest were included in linear models as either independent variables or covariates, where individual residualized probe M-values were included as dependent variables, while controlling for employment/disability status. Importantly, only exposures with a fully-mediating 5mC cluster were considered independent variables of interest. Other exposures which did not exhibit a fully-mediating 5mC cluster were considered as covariates. For probes/ASEs with nominally significant relationships (p < 0.05), we extracted probe p-values and probe names and used them as input to gene set enrichment analyses (GSEA) with the “methylGSA” package[94]. Importantly, ASE-associated probe p-values were only captured for ASEs in which a full mediator was previously observed, which limited analyses to CTQ only. GO sets of 25 to 1,000 genes were allowed, eliminating high-level GO-terms such as “biological process”, which facilitated testing of 5,478 gene sets. To produce a condensed summary of non-redundant GO-terms, the web-based tool “Revigo” was used[95].

## Results

### Study participants

Descriptive statistics for demographic and psychosocial measures in study participants are shown in Table 1. Thirty percent of the sample reported household income in the range of $0 to $499 per month, while 34% of the sample reported household income above $1000 per month. Fifteen percent of the sample reported not having finished high school, whereas 55% of the sample reported having more schooling than high school. Mean CTQ total value for participants was 40.5 (+/- 15.4), whereas mean TEI value for participants was 4.2 (+/- 2.3). Pearson correlations between variables used in the current study are shown in Figure 2.

**Table 1.** Demographic & psychosocial measure summary statistics (non-scaled) for the current sample (n = 97)

| Measure | Description | Value |
| --- | --- | --- |
| Age | Mean [SD] (Range) | 40 [12.5] (19 - 62) |
| Sex | Female | 100% |
| Self-reported race/ethnicity | African American | 100% |
| Household income (income) | \$0-499 | 30% |
| | \$499-999 | 36% |
| | \$1000+ | 34% |
| Educational attainment (education) | Some high school | 15% |
|  | High school grad or GED | 30% |
|  | ≥ Undergraduate degree | 55% |
| Employment/disability | Employed | 33% |
|  | Disabled (not employed) | 7% |
|  | Not employed or disabled | 60% |
| Childhood trauma questionnaire (CTQ total) | Mean [SD] (Range) | 40.5 [15.4] (25 - 93) |
| Traumatic events inventory (TEI) | Mean [SD] (Range) | 4.2 [2.3] (0 - 10.7) |
| Legend: [standard deviation], (range). |  |  |

**Figure 2.**
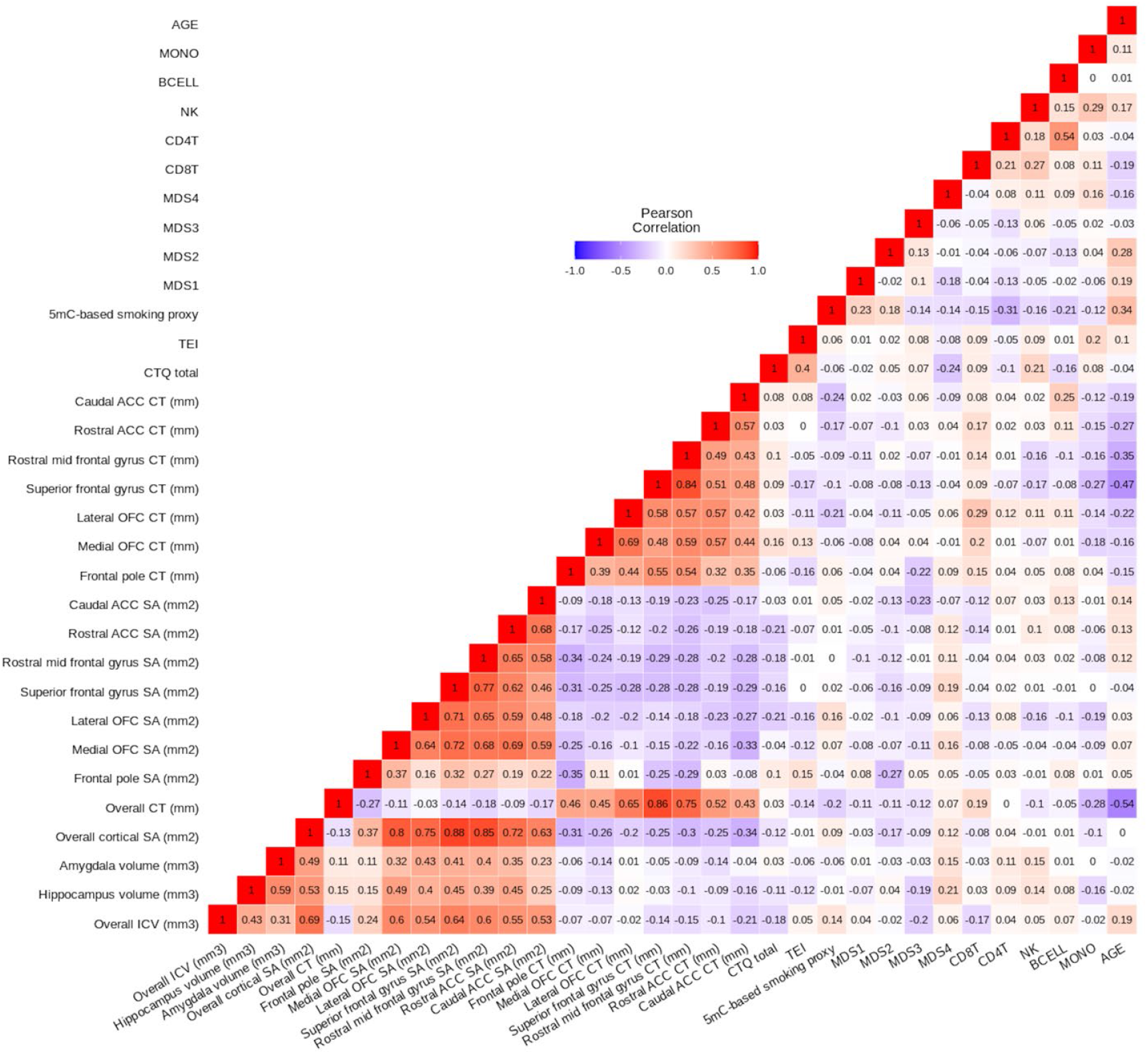
Pearson’s correlation heatmap of variables used throughout the current study. Age is negatively associated with multiple frontal cortex cortical thickness (CT) measures (Pearson’s correlation r_range_ = -0.15 to -0.47, p_range_ = 0.092 to 6×10^-7^). Within frontal cortex CT measures, multiple subregions show positive correlations with one another (Pearson’s correlation r_range_ = 0.32 to 0.84, p_range_ = 0.004 to < 2×10^-16^). The same phenomenon is observed within frontal cortex surface area (SA) measures (Pearson’s correlation r_range_ = 0.16 to 0.77, p_range_ = 0.210 to <2×10^-16^). Overall CT is positively correlated with each of the frontal cortex CT measures (Pearson’s correlation r_range_ = 0.43 to 0.86, p_range_ = 1×10^-7^ to <2×10^-16^), and overall SA is positively correlated with each of the frontal cortex SA measures (Pearson’s correlation r_range_ = 0.37 to 0.88, p_range_ = 5×10^-4^ to < 2×10^-16^).

### ASEs predict fronto-limbic brain morphometry

At nominal significance, CTQ total was negatively associated with RMFG SA (β = - 0.231, SE = 0.111, t = - 2.079, p = 0.041), but was not associated with other outcomes of interest (p > 0.05). Relative to high educational attainment (completed more schooling than high school), low educational attainment (no high school certificate) was negatively associated with cACC SA (β = - 0.855, SE = 0.280, t = - 3.049, p = 0.003), and positively associated with FP CT (β = 0.684, SE = 0.302, t = 2.267, p = 0.026) and SFG CT (β = 0.553, SE = 0.256, t = 2.162, p = 0.034) at the nominal significance threshold, but no other brain morphometry outcomes of interest (p > 0.05). On the other hand, relative to high income ($1000+/month), low income ($0-$499/month) was nominally negatively associated with amygdala GMV (β = - 0.649, SE = 0.283, t = - 2.299, p = 0.024), but no other neuroimaging outcome of interest (p > 0.05). TEI was not associated with any neuroimaging measures (p > 0.05). Nominally significant model outcomes can be found in Table 2. In controlling for 64 total tests at FDR = 0.10, no outcomes were BH-significant. Brain morphometry outcomes nominally associated with ASEs were carried into downstream analyses.

**Table 2.** Adverse social exposures (ASEs) predict hemisphere-mean fronto-limbic brain morphometry.

|  | Low household income |  |  |  | Low educational attainment |  |  |  | Childhood trauma burden |  |  |  | Adult trauma burden |  |  |  |
| --- | --- | --- | --- | --- | --- | --- | --- | --- | --- | --- | --- | --- | --- | --- | --- | --- |
| | $\beta$ | SE | T | P | $\beta$ | SE | T | P | $\beta$ | SE | T | P | $\beta$ | SE | T | P |
| Hipp. Volume (mm3) | -0.425 | 0.267 | -1.590 | 0.116 | 0.005 | 0.279 | 0.019 | 0.985 | 0.030 | 0.115 | 0.260 | 0.796 | -0.167 | 0.101 | -1.644 | 0.104 |
| Amyg. Volume (mm3) | -0.649 | 0.283 | -2.299 | 0.024* | 0.091 | 0.294 | 0.310 | 0.757 | 0.145 | 0.118 | 1.228 | 0.223 | -0.146 | 0.106 | -1.380 | 0.171 |
| FP SA (mm2) | 0.556 | 0.285 | 1.950 | 0.055 | -0.065 | 0.294 | -0.221 | 0.826 | -0.004 | 0.114 | -0.037 | 0.971 | 0.109 | 0.107 | 1.017 | 0.312 |
| Medial OFC SA (mm2) | 0.271 | 0.293 | 0.926 | 0.357 | -0.374 | 0.302 | -1.237 | 0.220 | -0.052 | 0.117 | -0.440 | 0.661 | -0.112 | 0.110 | -1.021 | 0.310 |
| Lateral OFC SA (mm2) | -0.033 | 0.298 | -0.111 | 0.912 | -0.244 | 0.307 | -0.796 | 0.428 | -0.211 | 0.119 | -1.769 | 0.081 | -0.093 | 0.111 | -0.834 | 0.407 |
| SFG SA (mm2) | 0.386 | 0.289 | 1.336 | 0.185 | -0.435 | 0.298 | -1.460 | 0.148 | -0.223 | 0.116 | -1.926 | 0.058 | 0.039 | 0.108 | 0.358 | 0.721 |
| RMFG SA (mm2) | -0.050 | 0.278 | -0.181 | 0.857 | -0.543 | 0.286 | -1.897 | 0.061 | -0.231 | 0.111 | -2.079 | 0.041* | 0.036 | 0.104 | 0.346 | 0.730 |
| Rostral ACC SA (mm2) | -0.176 | 0.291 | -0.602 | 0.549 | -0.511 | 0.301 | -1.700 | 0.093 | -0.229 | 0.117 | -1.961 | 0.053 | 0.029 | 0.109 | 0.268 | 0.790 |
| Caudal ACC SA (mm2) | 0.121 | 0.272 | 0.443 | 0.659 | -0.855 | 0.280 | -3.049 | 0.003** | -0.110 | 0.109 | -1.006 | 0.317 | 0.044 | 0.102 | 0.436 | 0.664 |
| FP CT (mm) | -0.137 | 0.293 | -0.468 | 0.641 | 0.684 | 0.302 | 2.267 | 0.026* | -0.079 | 0.117 | -0.671 | 0.504 | -0.120 | 0.110 | -1.098 | 0.275 |
| Medial OFC CT (mm) | -0.002 | 0.293 | -0.008 | 0.993 | 0.346 | 0.315 | 1.099 | 0.275 | 0.138 | 0.118 | 1.172 | 0.244 | 0.022 | 0.111 | 0.202 | 0.840 |
| Lateral OFC CT (mm) | -0.023 | 0.304 | -0.074 | 0.941 | 0.241 | 0.314 | 0.769 | 0.444 | 0.055 | 0.122 | 0.451 | 0.653 | -0.112 | 0.114 | -0.983 | 0.328 |
| SFG CT (mm) | -0.088 | 0.248 | -0.356 | 0.723 | 0.553 | 0.256 | 2.162 | 0.034* | 0.070 | 0.100 | 0.699 | 0.487 | -0.170 | 0.093 | -1.831 | 0.071 |
| RMFG CT (mm) | -0.078 | 0.281 | -0.276 | 0.783 | 0.443 | 0.290 | 1.530 | 0.130 | 0.072 | 0.113 | 0.637 | 0.526 | -0.112 | 0.105 | -1.062 | 0.291 |
| Rostral ACC CT (mm) | -0.063 | 0.297 | -0.211 | 0.833 | 0.060 | 0.318 | 0.188 | 0.851 | 0.019 | 0.118 | 0.163 | 0.871 | -0.034 | 0.112 | -0.305 | 0.761 |
| Caudal ACC CT (mm) | -0.124 | 0.296 | -0.419 | 0.676 | 0.187 | 0.318 | 0.589 | 0.558 | 0.046 | 0.119 | 0.388 | 0.699 | 0.004 | 0.112 | 0.039 | 0.969 |

\*\*\* P < 0.001, \*\*P < 0.01, \* P < 0.05, **P: BH significant (bold)**
Both dependent and independent variables were scaled prior to analysis. Childhood trauma burden was measured with the Childhood Trauma Questionnaire, and adult trauma burden was measured with the Traumatic Events Inventory. In separate linear models including hippocampus and amygdala volumes as dependent variables, income, education, CTQ total, and TEI were included as independent variables of interest, whereas genomic ancestry, age, employment/disability and ICV were included as covariates. The same independent variables and covariates were included, minus ICV, to predict frontal cortex subregions SA and CT measures.
Abbreviations. Hipp: hippocampus, Amyg: amygdala, FP: frontal pole, mOFC: medial orbito-frontal cortex, IOFC: lateral orbito-frontal cortex, SFG: superior frontal gyrus, RMFG: rostral medial frontal gyrus, rACC: rostral anterior cingulate cortex, cACC: caudal anterior cingulate cortex, SA: surface area, CT: cortical thickness, $\beta$ : scaled beta value, SE: standard error, T: t statistic value.

### ASEs predict 5mC ME

CTQ total was nominally associated with seven MEs, the strongest of which were Darkolivegreen (Darkolivegreen β = 0.354, SE = 0.110, t = 3.214, p = 0.002) and Steelblue (Steelblue β = - 0.306, SE = 0.111, t = - 2.748, p = 0.007). Compared to high educational attainment, low educational attainment was positively associated with one ME (Coral1 β = 0.750, SE = 0.286, t = 2.618, p = 0.010), and negatively associated with one ME (Lightcoral β = - 0.615, SE = 0.296, t = - 2.079, p = 0.040) at nominal levels. Relative to high income ($1000+/month), low income ($0-$499/month) was positively associated with three MEs (Antiquewhite4 β = 0.770, SE = 0.277, t = 2.781, p = 0.007; Indianred4 β = 0.717, SE = 0.276, t = 2.602, p = 0.011; Orange β = 0.571, SE = 0.274, t = 2.084, p = 0.040) at nominal levels. Because TEI was not associated with any fronto-limbic brain morphometric measures (Figure 1, Arm A), we do not report any results for this exposure. Results from all Arm B analyses can be found in Supplementary Table 1. In controlling for 73 tests at FDR = 0.10, no outcomes were BH-significant. 5mC MEs nominally associated with ASEs were carried into downstream analyses.

### 5mC MEs predict fronto-limbic brain morphometry

Four out of seven CTQ-associated MEs showed a nominally significant relationship with hemisphere-mean RMFG SA (Maroon, White, Tan, Yellow). The three strongest relationships were the Maroon ME (β = - 0.451, SE = 0.091, t = - 4.966, p = 3×10^-6^) (Figure 3a), White ME (β = - 0.392, SE = 0.093, t = - 4.231, p = 6×10^-5^) (Figure 3b), and Tan ME (β = 0.233, SE = 0.100, t = 2.322, p = 0.023) (Figure 3c). Two low educational attainment-associated MEs were tested independently for associations with hemisphere-mean cACC SA, frontal pole CT, and SFG CT. Of these, the Lightcoral ME was nominally associated with cACC SA (β = 0.215, SE = 0.099, t = 2.182, p = 0.032) (Figure 3d). Finally, of the three low income-associated MEs, none showed a nominally significant relationship with hemisphere-mean amygdala volume (Supplementary Table 2). In controlling for 16 tests at FDR = 0.10, the Maroon and White ME relationships with hemisphere-mean RMFG SA were BH-significant. 5mC MEs nominally associated with fronto-limbic brain morphometry measures were carried into downstream analyses.

**Figure 3.**
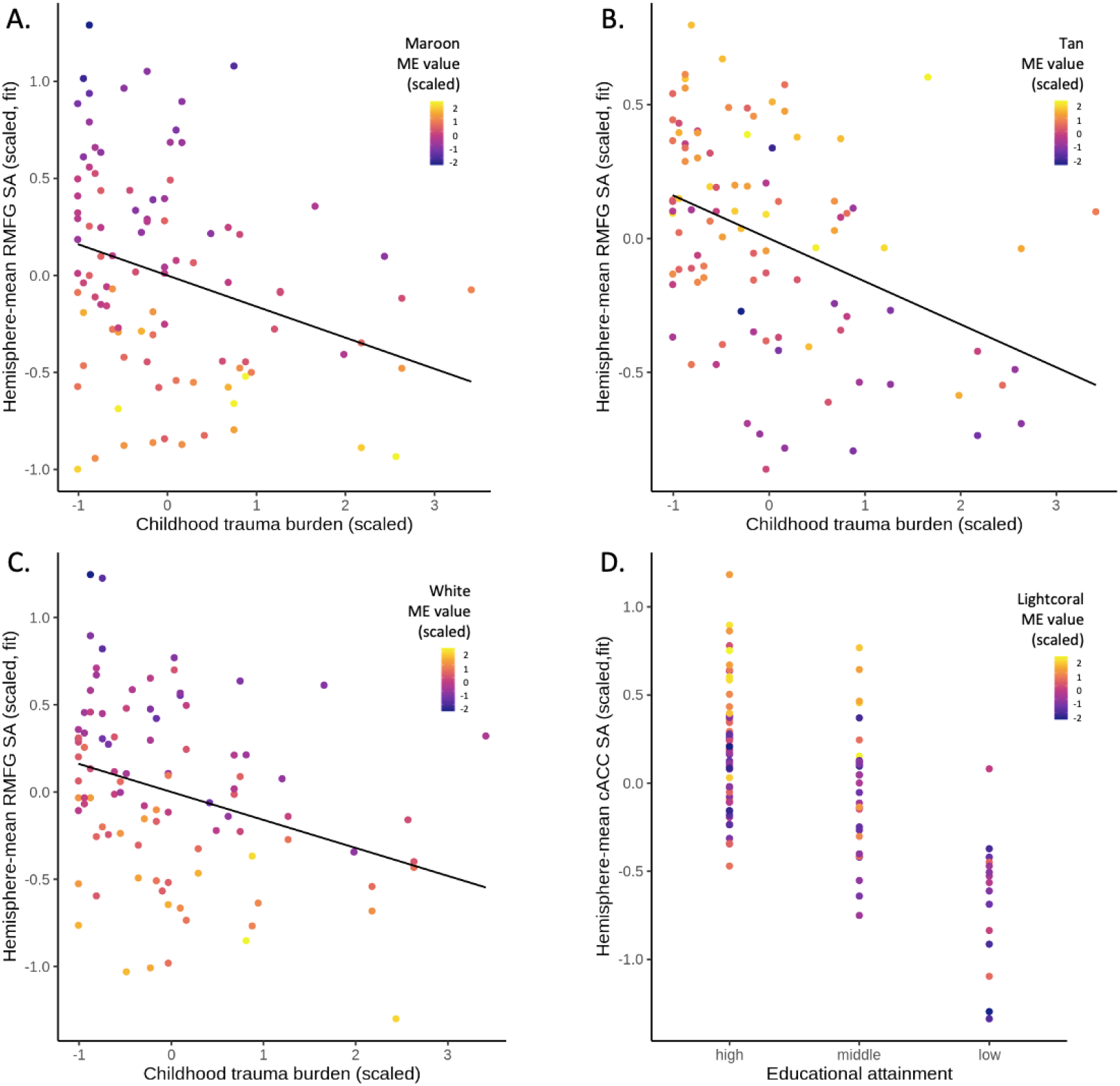
CTQ total is negatively associated with rostral middle frontal gyrus (RMFG) surface area (SA). Low educational attainment status is associated with low caudal anterior cingulate cortex (cACC) SA. Out of seven CTQ-associated module eigengenes (MEs), the three strongest relationships with hemisphere-mean RMFG SA were the Maroon (A), White (B), and Tan (C) MEs. Out of two low education attainment-associated MEs, the strongest relationship with hemisphere-mean cACC SA was the Lightcoral ME (D). As shown, neuroimaging measures are adjusted by age, ME value, genomic ancestry, employment/disability status, and an adverse social exposure (ASE). Dependent and independent variables were scaled. A. CTQ total was positively associated with the Maroon ME (β = 0.289, SE = 0.109, t = 2.639, p = 9.8×10^-3^). In turn, the Maroon ME was negatively associated with RMFG SA (β = - 0.451, SE = 0.091, t = - 4.966, p = 3×10^-6^). B. CTQ total was negatively associated with the Tan ME (β = - 0.248, SE = 0.112, t = - 2.220, p = 0.029). The Tan ME was, in turn, positively associated with RMFG SA (β = 0.233, SE = 0.100, t = 2.322, p = 0.023). C. CTQ total was positively associated with the White ME (β = 0.236, SE = 0.110, t = 2.151, p = 0.034). In turn, the White ME was negatively associated with RMFG SA (β = - 0.392, SE = 0.093, t = - 4.231, p = 6×10^-5^). D. Low educational attainment (no high school certificate) was associated with the Lightcoral ME (β = - 0.615, SE = 0.296, t = - 2.079, p = 0.040). The Lightcoral ME, in turn, was positively associated with cACC SA (β = 0.215, SE = 0.099, t = 2.182, p = 0.032).

### ME mediation

Four CTQ-associated MEs that also showed a nominally significant relationship with hemisphere-mean RMFG SA were tested for their mediating status between CTQ total and hemisphere-mean RMFG SA (Maroon, White, Tan, Yellow). Both the Maroon ME (TE β = -0.983, p = 0.047; IDE β = -0.466, p = 0.008; DE β = -0.517, p = 0.277) and the White ME (TE β = -1.014, p = 0.034; IDE β = -0.348, p = 0.032; DE β = -0.667, p = 0.164) were full mediators of this relationship. TE results show that RMFG SA was an estimated ∼589 mm^2^ lower at the highest levels of CTQ exposure compared to the lowest levels of CTQ exposure. Independently, the Maroon ME accounted for 275 mm^2^ (47%) of that effect, whereas the White ME accounted for 204 mm^2^ (34%). Neither the Tan nor Yellow MEs were partial or full mediators of the relationship between CTQ and hemisphere-mean RMFG SA (TE p < 0.05; IDE p > 0.05; DE p > 0.05), although the direction of TEs in these analyses closely mirrored those in the Maroon and White ME mediation analyses. The Lightcoral ME, which we previously observed to be associated with low education attainment and cACC SA, was neither a partial nor full mediator of the relationship between low education attainment and hemisphere-mean cACC SA (TE β = -0.844, p = 0.003; IDE β = -0.107, p = 0.100; DE β = - 0.737, p = 0.010). However, the DE indicates that low educational attainment status accounted for cACC SA that is 84 mm^2^ smaller than high educational attainment status. In controlling for ten tests of the IDE and DE p-values for the five relationships above at FDR = 0.10, all nominally significant ME IDEs were BH-significant and all DEs remained non-significant (Table 3). Follow-up mediation analyses were then performed on individual probes from the Maroon and White modules.

**Table 3.**
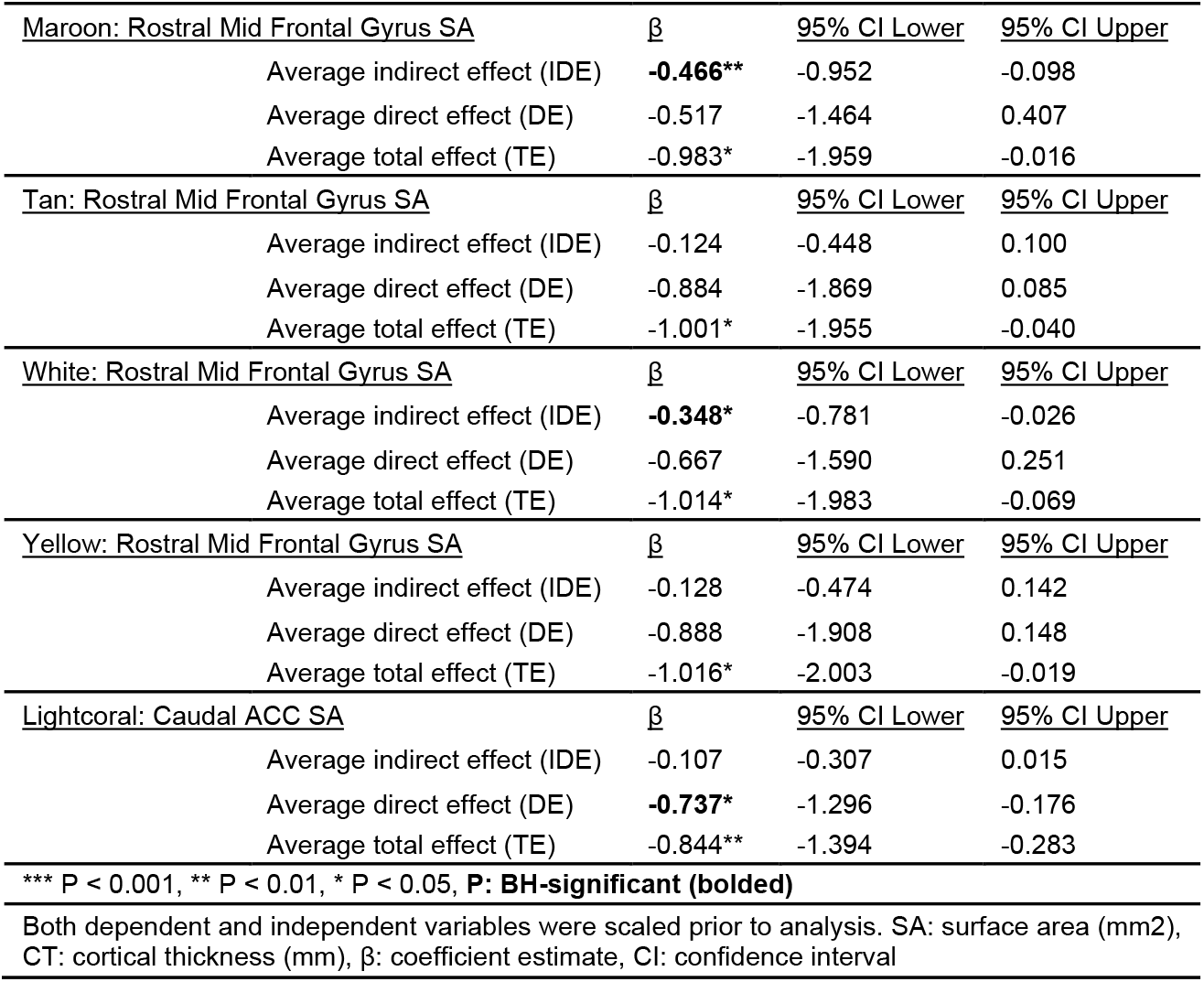
Module eigengenes (MEs) mediating observed adverse social exposure (ASE), fronto-limbic brain morphometry relationships.

| <u>Maroon: Rostral Mid Frontal Gyrus SA</u> | <u>$\beta$</u> | <u>95% CI Lower</u> | <u>95% CI Upper</u> |
| --- | --- | --- | --- |
| Average indirect effect (IDE) | **-0.466\*\*** | -0.952 | -0.098 |
| Average direct effect (DE) | -0.517 | -1.464 | 0.407 |
| Average total effect (TE) | -0.983* | -1.959 | -0.016 |
| <u>Tan: Rostral Mid Frontal Gyrus SA</u> | <u>$\beta$</u> | <u>95% CI Lower</u> | <u>95% CI Upper</u> |
| Average indirect effect (IDE) | -0.124 | -0.448 | 0.100 |
| Average direct effect (DE) | -0.884 | -1.869 | 0.085 |
| Average total effect (TE) | -1.001* | -1.955 | -0.040 |
| <u>White: Rostral Mid Frontal Gyrus SA</u> | <u>$\beta$</u> | <u>95% CI Lower</u> | <u>95% CI Upper</u> |
| Average indirect effect (IDE) | **-0.348\*** | -0.781 | -0.026 |
| Average direct effect (DE) | -0.667 | -1.590 | 0.251 |
| Average total effect (TE) | -1.014* | -1.983 | -0.069 |
| <u>Yellow: Rostral Mid Frontal Gyrus SA</u> | <u>$\beta$</u> | <u>95% CI Lower</u> | <u>95% CI Upper</u> |
| Average indirect effect (IDE) | -0.128 | -0.474 | 0.142 |
| Average direct effect (DE) | -0.888 | -1.908 | 0.148 |
| Average total effect (TE) | -1.016* | -2.003 | -0.019 |
| <u>Lightcoral: Caudal ACC SA</u> | <u>$\beta$</u> | <u>95% CI Lower</u> | <u>95% CI Upper</u> |
| Average indirect effect (IDE) | -0.107 | -0.307 | 0.015 |
| Average direct effect (DE) | **-0.737\*** | -1.296 | -0.176 |
| Average total effect (TE) | -0.844** | -1.394 | -0.283 |

\*\*\* P < 0.001, \*\* P < 0.01, \* P < 0.05, **P: BH-significant (bolded)**
Both dependent and independent variables were scaled prior to analysis. SA: surface area (mm<sup>2</sup>), CT: cortical thickness (mm), $\beta$ : coefficient estimate, CI: confidence interval

### Probe-wise mediation: Maroon module

Six out of the 11 Maroon ME probes were full mediators of the relationship between CTQ total and RMFG SA (Table 4). The top three strongest mediators based on IDE β coefficients were: cg21622733 (TE β = -0.983, p = 0.044; IDE β = -0.512, p = 0.003; DE β = -0.471, p = 0.320), cg19805668 (TE β = -1.003, p = 0.042; IDE β = -0.411, p = 0.009; DE β = -0.592, p = 0.217), and cg03710029 (TE β = -0.990, p = 0.048; IDE β = -0.321, p = 0.019; DE β = -0.670, p = 0.182). Respectively, these probes mapped to the long intergenic non-coding (LINC) RNA gene *LINC01531*, the placental growth factor gene *PGF*, and the solute carrier family 38 member 10 gene *SLC38A10*. The other three full mediating probes mapped to myosin heavy chain 14 (*MYH14*), DNA methyltransferase 3 alpha (*DNMT3A*), and neuralized E3 ubiquitin protein ligase 4 (*NEURL4)*. In controlling for 22 tests of Maroon probe IDE and DE p-values, only two nominally significant probes were also BH-significant (cg21622733: *LINC01531* and cg19805668: *PGF*). Comprehensive probe-wise mediation results are found in Supplementary Table 3.

**Table 4.** Probe-wise mediation analysis of the relationship between childhood trauma burden (CTQ total) & rostral middle frontal gyrus (RMFG) surface area (SA)

| Probe name | Module name | Chr:pos (hg38) | Gene name | IDE $\beta$ | IDE P | DE $\beta$ | DE P | TE $\beta$ | TE P |
| --- | --- | --- | --- | --- | --- | --- | --- | --- | --- |
| **cg21622733** | **Maroon** | Chr19:35408551 | *LINC01531* | **-0.512** | **0.003\*\*** | -0.471 | 0.320 | -0.983 | 0.044* |
| **cg19805668** | **Maroon** | Chr14:74951972 | *PGF* | **-0.411** | **0.009\*\*** | -0.592 | 0.217 | -1.003 | 0.042* |
| cg10592766 | Maroon | Chr19:50226846 | *MYH14* | -0.321 | 0.019* | -0.670 | 0.182 | -0.990 | 0.048* |
| cg03710029 | Maroon | Chr17:81291801 | *SLC38A10* | -0.383 | 0.025* | -0.613 | 0.188 | -0.996 | 0.043* |
| cg15150970 | Maroon | Chr2:25250660 | *DNMT3A* | -0.253 | 0.049* | -0.753 | 0.125 | -1.006 | 0.040* |
| cg12219789 | Maroon | Chr17:7324180 | *NEURL4* | -0.352 | 0.049* | -0.622 | 0.183 | -0.974 | 0.049* |
| cg01544227 | White | Chr12:8323720 | *AC092745.5* | -0.305 | 0.036* | -0.689 | 0.175 | -0.994 | 0.044* |
| cg00686169 | White | Chr8:8024026 | *FAM90A24P* | -0.277 | 0.043* | -0.740 | 0.126 | -1.017 | 0.037* |
| cg08002107 | White | Chr8:12135255 | *USP17L7* | -0.268 | 0.047* | -0.738 | 0.122 | -1.006 | 0.040* |

\*\*\* P < 0.001, \*\*P < 0.01, \* P < 0.05, **P: BH-significant (bold)**
Both dependent and independent variables were scaled prior to analysis. IDE: indirect effect, DE: direct effect, TE: total effect, $\beta$ : coefficient estimate, P: nominal p-value, BH: Benjamini-Hochberg

In all, probes from the Maroon module map to eight known protein coding genes (*MYH14, HGSNAT, SFTPA1, PGF, NEURL4, DNMT3A, SLC38A10*, and *PEBP4*) and two non-coding RNAs (*LINC01531 and MIR3659HG*). GO-terms associated with these loci include: *MYH14* (neuronal action potential GO:0019228, actomyosin structure organization GO:0031032), *HGSNAT* (lysosomal transport GO:0007041, neutrophil degranulation GO:0043312), *SFTPA1* (toll-like receptor signaling pathway GO:0002224), *PGF* (chemoattractant activity GO:0042056, vascular endothelial growth factor receptor binding GO:0005172), *NEURL4* (ubiquitin protein ligase activity GO:0061630), *DNMT3A* (DNA (cytosine-5-methyltransferase activity GO:0003886), *SLC38A10* (amino acid transmembrane transporter activity GO:0015171), and *PEBP4* (protein binding GO:0005515), among others.

### Probe-wise mediation: White module

Three out of 19 White ME probes were full mediators of the relationship between CTQ total and RMFG SA (Table 4): cg01544227 (TE β = -0.994, p = 0.044; IDE β = -0.305, p = 0.036; DE β = -0.689, p = 0.175), cg00686169 (TE β = -1.017, p = 0.037; IDE β = -0.277, p = 0.043; DE β = -0.740, p = 0.126), and cg08002107 (TE β = -1.006, p = 0.040; IDE β = -0.268, p = 0.047; DE β = -0.738, p = 0.122). These three probes mapped to *AC092745*.*5, FAM90A24P*, and *USP17L7* loci, respectively. In controlling for 38 tests of White probe IDE and DE p-values, no nominally significant probes were BH-significant. Comprehensive probe-wise mediation results are found in Supplementary Table 3.

In all, only one probe from the White module maps to a known protein coding gene, *USP17L7*, which is associated with the thiol-dependent de-ubiquitinase GO term (GO:0004843). One other probe maps to a LINC RNA, *FAM66D*. The remaining probes map to a pseudogene family, named family with sequence similarity 90. Members include: *FAM90A10, FAM90A11P, FAM90A20, FAM90A24P, FAM90A25P, FAM90A5, FAM90A6P*, and *FAM90A7*. The only GO term associated with these pseudogenes is nucleic acid binding (GO:0003676).

### Gene set enrichment analysis

CTQ total was included as the independent variable of interest in linear models with probe residualized M-values as dependent variables. Income, education, and TEI were included and considered as covariates, as no 5mC clusters mediated the relationships between our exposures and outcomes of interest. We used the resultant 6,335 nominally significant p-values and probe names as GSEA input, facilitating the testing of 5,478 GO-terms. After Revigo redundancy reduction, 30 BH-significant GO-terms remained for interpretation. These included immune-related GO-terms: macrophage migration (GO:1905517, p = 9×10^-8^, rank = 10), T cell receptor complex (GO:0042101, p = 6×10^-6^, rank = 13), and chemokine-mediated signaling pathway (GO:0070098, p = 7×10^-4^, rank = 28), among four others. Two CNS-related GO-terms were present: negative regulation of smoothened signaling pathway (GO:0045879, p = 2×10^-6^, rank = 12), and glutamine family amino acid biosynthetic process (GO:0009084, p = 1×10^-4^, rank = 19), along with one HPA-axis related term: thyroid hormone metabolic process (GO:0042403, p = 2×10^-15^, rank = 5). Additionally, numerous signal transduction and membrane transport GO-terms were present: positive regulation of phospholipase activity (GO:0010518, p = 5×10^-25^, rank = 1), intrinsic component of peroxisomal membrane (GO:0031231, p = 7×10^-18^, rank = 2), and RNA polymerase II carboxy-terminal domain kinase activity (GO:0008353, p = 8×10^-15^, rank = 6), among others (Supplementary Table 4).

## Discussion

Here we applied genome-scale approaches to a sample of African American women from a low income, highly trauma exposed environment, representing an understudied population, where our main goal was to assess whether blood-derived 5mC cluster MEs might index CNS endophenotypes of stress-related psychiatric illness. Our study findings indicate that high childhood trauma burden, low educational attainment, and low income are each associated with CNS endophenotypes of stress-related psychiatric illness, and that a subset of blood-derived 5mC MEs statistically mediate the relationship between childhood trauma burden and RMFG SA. We further found that the individual probes from mediating 5mC MEs fell in genes with CNS-relevant and immune system GO-functions. Finally, we found that the underlying childhood trauma burden-associated methylomic network was enriched with HPA-axis, immune, and CNS-related gene sets, in addition to signal transduction and membrane transport functions. Overall, we posit that the peripheral epigenetic signatures mediating our relationships of interest are consistent with previously observed patterns in which stress-related psychiatric illness is accompanied by profiles of peripheral inflammation[31, 96-98].

In the current study, childhood trauma burden was negatively associated with hemisphere-mean RMFG SA, which mirrors findings from cohorts enriched for stress, trauma, and psychiatric illness[99, 100]. We also observed higher CT of both the FP and SFG in participants with low educational attainment, as well as higher cACC SA. Although studies on FP CT and ASEs (besides trauma) are somewhat limited, our findings support past research investigating whether high CT is representative of a predisposition to psychiatric illness development, or whether high CT emerges after psychiatric illness onset. Specifically, prior longitudinal work showed higher cortical thickness in participants at high risk of mood disorder compared to healthy controls at baseline; the high-risk participants who went on to develop MDD then showed increased FP CT over the two-year time period following their diagnosis[101]. This research implies that high FP CT could represent both a predisposition to MDD development risk, and a post-onset emergent phenotype of MDD.

Our study also showed that low household income was negatively associated with hemisphere-mean amygdala GMV. This finding aligns with research in children/adolescents showing that low familial income-to-needs ratio is associated with decreased amygdala GMV [92], and with studies in adults showing that past year financial hardship is associated with decreased amygdala GMV [102]. However, the relationships between socioeconomic status-related measures and amygdala GMV are well-studied, and variable conclusions between studies are common[103]. Hemisphere-mean hippocampal volume was not associated with any ASE; as in the case of amygdala GMV, study conclusions are mixed[103].

In all, four blood-derived MEs were tested for mediating status between childhood trauma burden and RMFG SA. Both the Maroon and the White ME fully mediated the relationship and accounted for significant proportions of variability (47% and 35% respectively). The Maroon module is composed of 11 probes, six of which are also full mediators of the aforementioned relationship. The six probes fully mediating the childhood trauma burden and RMFG SA relationship are located in the *MYH14, PGF, NEURL4, DNMT3A, SLC38A10*, and *LINC01531*. GO-terms associated with these loci include: *MYH14* (neuronal action potential GO:0019228, actomyosin structure organization GO:0031032), *PGF* (chemoattractant activity GO:0042056, vascular endothelial growth factor receptor binding GO:0005172), *NEURL4* (ubiquitin protein ligase activity GO:0061630), *DNMT3A* (DNA (cytosine-5-methyltransferase activity GO:0003886), and *SLC38A10* (amino acid transmembrane transporter activity GO:0015171). Of particular interest is the probe falling in the *DNMT3A* gene. Past longitudinal study in a predominantly female African American sample showed differential blood *DNMT3A* 5mC in response to traumatic event exposure[104], and emerging work focused specifically on African ancestry individuals has identified a genome-wide significant association in *DNMT3A* in relation to MDD[105]. What is more, in a rodent model of fear conditioning, *Dnmt3a* mRNA expression is altered in forebrain neurons, leading to differential DNA 5mC of synaptic plasticity and memory formation genes[106]. *Dnmt3A* mRNA expression in rodent nucleus accumbens is also altered in response to chronic restraint stress, and targeted inhibition of *Dnmt3A* 5mC potentiates anti-depressant-like effects[107]. Based on the current and prior findings, it appears that the *DNMT3A* gene in humans is a potential locus of stress- or trauma-related biological embedding, and that measurements of *DNMT3A* 5mC taken from peripheral tissues could be indexing fronto-limbic variability associated with these exposures. Overall, our findings in the Maroon module imply that immune signaling, cellular proliferation, neuronal development, and epigenetic regulatory pathways could be potential substrates for the biological embedding of childhood trauma.

The second fully mediating ME was the White module comprised of 19 probes, three of which were full mediators of the relation between CTQ total and RMFG SA. Thirteen probes from this module are mapped to the “Family with sequence similarity 90” superfamily of pseudogenes, and all but two probes exist in a one megabase stretch of chromosome eight. The three full mediator probes (cg00686169, cg01544227, and cg08002107) fall in the *FAM90A24P, LINC00937, and FAM66D* loci, respectively. Little is known of the *FAM90A24P* locus, but the *LINC00937* locus encodes a LINC RNA species that is strongly associated with cutaneous melanoma prognosis[108], as well as endocervical cancer progression[109], suggesting that it plays a role in cellular proliferation. On the other hand, the *FAM66D* locus encodes a LINC RNA species whose expression is significantly upregulated in both Crohn’s disease (CD) and ulcerative colitis (UC), the two most common types of inflammatory bowel disease (IBD)[110]. Efforts made towards elucidating the function of the broader chromosome eight locus highlight its structurally dynamic nature, in addition to its harboring of the defensin genes, the protein products of which play significant roles in the innate immune response, as well as antitumor response[111]. Defensin proteins are also recognized as key contributors to innate immune defense against UC, CD, and IBD in general[112].

Taken together, results suggest that probes mediating the childhood trauma burden and RMFG SA relationship interact with endothelial growth and innate immune response regulatory pathways. This notion is supported by repeated observation of increased blood-based inflammation profiles, as measured in adults that had previously experienced childhood trauma exposure [113-115]. More CNS-focused research has also shown significant dysregulation of stress-response and inflammation-related mRNA/protein levels in post-mortem frontal cortex of patients with neurodevelopmental[116] and psychiatric illnesses[117, 118]. What is more, decreased CT of medial pre-frontal cortex is observed in MDD patients, a relationship that was partially mediated by inflammatory factors known to influence neuroplasticity[119]. However, the relevant studies informing our current results are sparse, and could be considered underpowered. Thus, while suggestive, more work remains to be done in order to draw robust inferences regarding the relationship between peripheral inflammation and neural endophenotypes of stress-related psychiatric illness.

In our GSEA of childhood trauma burden -associated probes, the top enriched term was “positive regulation of phospholipase activity”, meaning that multiple probes fall in genes that increase the frequency, rate, or extent of phospholipase activity. Of close relation is the 11^th^ ranked GO-term, “inositol phosphate phosphatase activity”. Together, these terms represent one of the major mechanisms of neuronal and hormonal signal transduction in mammals, the activation of phospholipase C (PLC) and subsequent activation of inositol signaling pathways[120]. In brief, extracellular stimuli receptor binding activates PLC. PLC then functions to convert phosphatidylinositol-4-5 bisphosphate (P45P2) into inositol-1-4-5 trisphosphate (I145P3). From this, the kinase-activating secondary messenger diacylglycerol (DAG) is activated[120]. There are many isoforms of the PLC enzyme family, but PLC-δ1 is strongly implicated in regulating cell-motility and cytoskeletal organization[121], and in inhibiting inflammatory immune responses[122]. Quantities of this PLC isoform, PLC-δ1, are highly correlated with nuclear levels of P45P2[123]; interestingly, P45P2 regulates transcription through binding interaction with the RNA polymerase II C-terminal tail[124]. The aforementioned processes are specifically represented within our GSEA results (Supplementary Table 4): RNA polymerase II carboxy-terminal domain kinase activity, regulation of mitotic spindle organization, and numerous immune signaling and motility terms. Given childhood trauma burden is the ASE implicated most strongly in our results, it stands to reason that in this sample of African American women, peripheral inflammatory signals could be heightened in those with high childhood trauma burden, and phospholipase activity may be upregulated in an attempt to restore homeostatic balance.

Overall, we posit that the blood-based epigenetic signatures mediating our relationships of interest are potentially explained by ASE-associated peripheral inflammation. In response to perceived stress or trauma, fronto-limbic brain regions associated with cognitive function, emotional reactivity, and memory are activated, signaling to the HPA-axis’ neural hub, and eventually potentiating glucocorticoid (GC) release. Critical to immunological, metabolic, cardiac, and homeostatic functions, GCs act mechanistically in and around the CNS and peripheral nervous system, including serving as a long-range negative feedback mechanism to inhibit further HPA-axis activity. Additionally, the ligands and receptors of cytokines and neurotransmitters are shared throughout the CNS, HPA-axis, and immune systems[125, 126]. Chronic stress signaling can result in neural growth, inflammation, metabolism, and stress-related pathway disruption through molecular and epigenetic mechanisms[40], which are consistent with 5mC-derived patterns observed in peripheral tissue in the current report.

The foremost strength of the current research is the inclusion of multi-modal data types (epigenetic/neuroimaging) to characterize the biological embedding of ASEs. Furthermore, the dimension reduction techniques used in the current work, clustering and principal component extraction, were valuable in reducing the burden of multiple hypothesis testing and in describing potentially related networks of 5mC loci. It also focused the analysis onto loci with greater prospect for proxy or surrogate status with etiologically-relevant CNS tissue. The composition of the sample is also a strength of the current work, as it reflects an understudied population at elevated risk for PTSD and for persistence of stress-related psychiatric illness[127]. However, because of the homogenous nature of the sample, we are unable to make confident conclusions about ancestry-specific or sex-specific effects. Limitations of the current study include relatively small sample size and the inability to infer causality from cross-sectional samples. In addition, the current research used peripheral tissue in lieu of etiological CNS tissue for obvious technical reasons.

Functional MRI measures such as neurotransmitter specific PET imaging could also provide deeper endophenotypic insights. Future studies on this topic should capture longitudinal data from a larger sample and could investigate genetic factors or tissues of etiological interest.

The current study showed that exposure to high childhood trauma burden, low educational attainment, and low income are each associated with neuroimaging endophenotypes of psychiatric illness, and that the relationship between childhood trauma and RFMG SA in particular is mediated by a subset of blood-derived 5mC measurements. In addition, we found that the mediating 5mC MEs were enriched with probes falling in genomic regulatory regions, and in genes with CNS-relevant and immune system GO-functions. Finally, we found that the underlying childhood trauma burden-associated methylomic network was enriched with HPA-axis, immune, and CNS-related gene sets. Together, these results highlight a feasible epigenetic mechanism through which ASEs become embedded in human physiological systems, and through which they contribute to the development of stress-related psychiatric illness. Although these concepts are broadly applicable across race, ethnicity, and biological sex, the current results highlight mechanisms of biological embedding that may be specific to African American women. These epigenetic signatures could be taken as peripheral biomarkers of perturbed underlying neurobiology associated with ASEs, and could further research efforts into etiological tissues of interest such as endocrine, immune, and CNS cell types. These findings could also serve to potentiate increased investigation of understudied populations at significant risk for onset or persistence of certain stress-related psychiatric illnesses.

## Data Availability

DNA methylation data analyzed in this paper are available at https://www.ncbi.nlm.nih.gov/geo/ (dataset GSE132203). Individual-level neuroimaging data will be made available to researchers following an approved analysis proposal through the GTP. For additional information on access to these data, including PI contact information for the GTP, please contact the corresponding author.

https://www.ncbi.nlm.nih.gov/geo/query/acc.cgi?acc=GSE132203

## Acknowledgements

This work was supported by the National Institutes of Mental Health (MH096764, MH071537, MH098212, MH111671, MH101380 and 2R01MH108826) and the National Institute of Minority Health and Health Disparities (2R01MD011728). Support was also received from Emory and Grady Memorial Hospital General Clinical Research Center, NIH National Centers for Research Resources (M01RR00039) and Howard Hughes Medical Institute.

